# Routine deworming during antenatal care decreases risk of neonatal mortality and low birthweight: a retrospective cohort of survey data

**DOI:** 10.1101/2020.04.07.20057000

**Authors:** Bhavneet Walia, Brittany L. Kmush, Sandra D. Lane, Timothy Endy, Antonio Montresor, David A. Larsen

## Abstract

**Background:** Soil transmitted helminths (STH) are a common infection among pregnant women in areas with poor access to sanitation. Deworming medications are cheap and safe; however, the health benefit of deworming during pregnancy is not clear.

**Methods / Principal Findings:** We created a retrospective cohort of more than 800,000 births from 95 Demographic and Health Survey datasets to estimate the impact of deworming medicine during routine antenatal care (ANC) on neonatal mortality and low birthweight. We first matched births on the probability of receiving deworming during ANC. We then modeled the birth outcomes with the matched group as a random intercept to estimate the effect of deworming during antenatal care after accounting for various risk factors. We also tested for effect modification of soil transmitted helminth prevalence on the impact of deworming during ANC. Receipt of deworming medication during ANC was associated with a 15% reduction in the risk of neonatal mortality (95% confidence interval = 11-18%, n = 797,772 births), with no difference between high and low transmission countries. In low transmission countries, we found an 11% reduction in the odds of low birth weight (95% confidence interval = 8-13%) for women receiving deworming medicine and in high transmission we found a 3% reduction in the odds of low birthweight (95% confidence interval = 1-5%).

**Conclusions / Significance:** These results suggest a substantial health benefit for deworming during ANC that may be even greater in countries with low STH transmission.

## Introduction

### Background/rationale

Soil transmitted helminths (STH) are parasitic nematodes (worms) that are transmitted by contamination of soil with human feces. The major species of STH that infect humans are *Ascaris lumbricoides* and *Trichuris trichiura* (whipworm), which infect humans via a fecal-oral route, and hookworm (*Anclostoma duodenale* and *Necator americanus*), whose eggs are shed in fecal matter and then hatched larvae burrow through the skin of the host. STH are estimated to infect more than two billion people across the globe [1], and in 2016 caused the loss of an estimated 3.5 million disability-adjusted life years (DALYs) [2].

STH affect human health in various ways. Hookworm is known to cause iron deficiency and anemia. For pregnant women, the resulting anemia can be particularly severe [3,4]. Infections with *T. trichiura* are also likely to cause anemia, and are associated with poor growth and delayed cognitive development in children [5]. Infections with *A. lumbricoides* are also associated with poor growth and delayed cognitive development in children [6].

Due to the fecal-oral route of transmission and the life stages in the soil, clean water and adequate sanitation access can easily prevent infections with STH [7–9]. However, sanitation access is still limited in lower-income countries, and therefore periodically clearing the parasites with deworming treatments is a short term intervention recommended by the WHO in STH endemic areas. The WHO manages a global donation of anthelminthics (albendazole and mebendazole) —with the support of several pharmaceutical companies who donate the medicines—providing them to endemic countries that request them for control programs targeting preschool children and school-age children. The effectiveness of deworming children has been called into question, however, due to recent reviews finding contradictory results of the health impact of mass drug administration in these populations [10–12]. A recent Cochrane review has also found limited evidence that deworming medicine during antenatal impacts birth outcomes or neonatal mortality [13]. However, the review evaluated less than 4,000 pregnancies in four studies, and the authors state that more data are needed to establish the benefit of the intervention or potential lack thereof.

### Objectives

In this study, we explore the connection between anthelminthic treatment of pregnant women during antenatal care and the outcomes of neonatal mortality and low-birth weight using a retrospective cohort of survey data of more than 770,000 births across a broad range of STH transmission settings.

## Methods

### Study design

We utilized birth histories from cross-sectional surveys to create a retrospective cohort to measure the impact of routine deworming medicine during antenatal care on subsequent neonatal mortality and low birthweight for births between 1998-2018 in 56 lower income countries (Figure 1).

**Figure 1:**
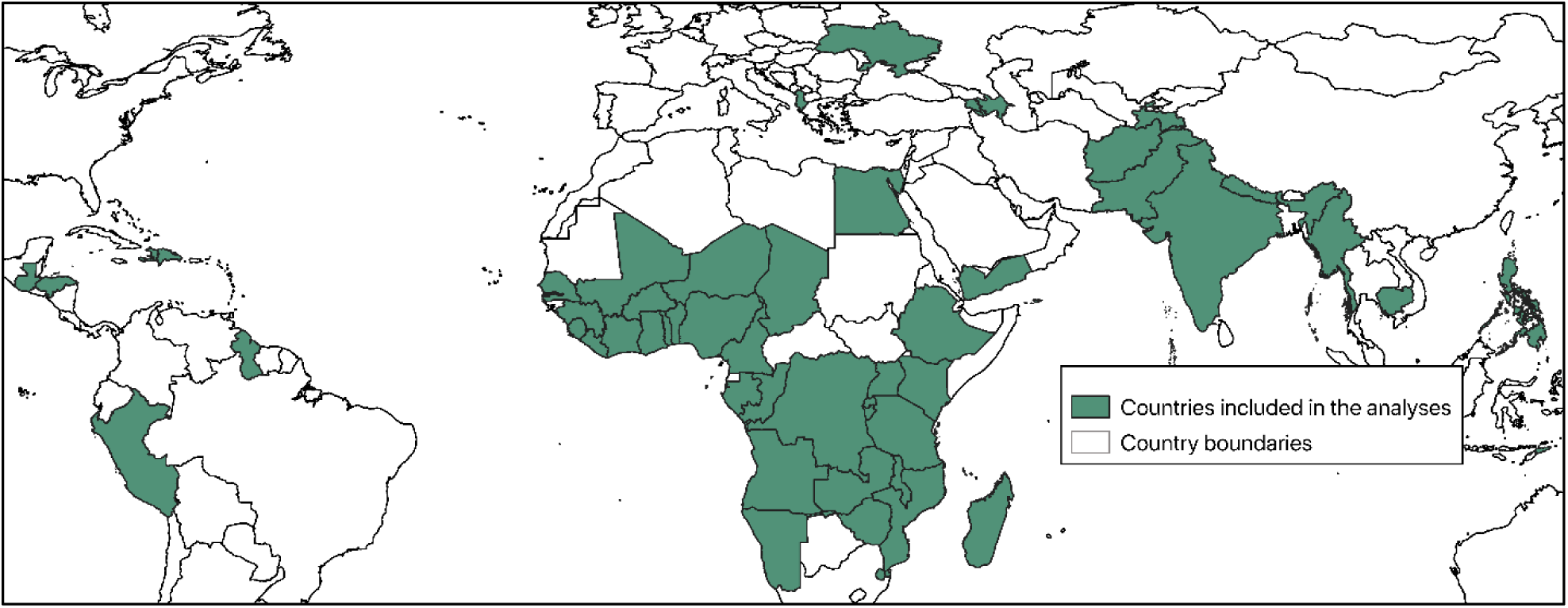
Map showing the countries included in the analyses.

### Data sources

The Demographic and Health Surveys (DHS) are funded at least in part by the United States Agency for International Development. These surveys utilize nationally representative two-stage cluster samples to generate information on child mortality and women’s fertility. As part of the survey, complete birth histories are recorded for all women aged 15-49 years of age, including whether or not each child is still alive and the age of the child’s death if the child died. Additionally, information on the most recent pregnancy within the previous 2 years is collected, including various aspects of the woman’s antenatal care (ANC) such as a question on the administration of deworming during the most recent pregnancy. We sought to include all DHS datasets with the following conditions: 1) the survey was conducted in 1990 or after, 2) the survey contained information on deworming during ANC, and 3) the survey was publicly available as of August 23, 2019.

### Outcomes

Neonatal mortality served as the primary outcome. In the DHS questionnaire, the survey respondent classifies the age of their child at death in terms of days, weeks, or months. Significant heaping of neonatal mortality occurs at one month of life in these data. We included children as neonatal deaths if their mother described them dying within the first 28 days of life, the first four weeks of life, or the first month of life.

Low birthweight served as the secondary outcome. In the DHS questionnaire, a child’s weight at birth is included if the child is weighed at birth (approximately 50% of babies in the DHS are weighed at birth). The mother is also asked the child’s perceived birth size. We created a composite indicator of measured low birthweight when available and perceived birth size when measured low birthweight was unavailable. For those children who were weighed at birth we categorized children as being low birth weight if they were < 2500 grams at birth. For those children who were not weighed at birth, we categorized children as low birth weight if they were perceived to be smaller than average or very small. We also conducted a sensitivity analysis wherein we limited the low birthweight analyses to those children who were weighed at birth.

### Bias

Women who attend routine antenatal care and receive deworming medicines are predisposed to have better birth outcomes than women who do not, independent of the effect of the deworming medicine. This may include, among other factors, better ANC as well as better post-natal care. We therefore utilized an exact matching procedure to pre-process the data and reduce the selection bias of receiving deworming medicine [14]. We exactly matched women on their probability of receiving deworming during pregnancy using the MatchIt package in R version 3.6.1 [15,16]. The following equation describes the matching process:

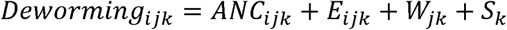

where *Deworming*_*ijk*_ is a dichotomous outcome for woman *i* in household *j* in survey *k*; *ANC*_*ijk*_ is a vector of various aspects of ANC, including being weighed during ANC, being measured during ANC, having blood pressure taken during ANC, giving a urine sample during ANC, giving a blood sample during ANC, receiving intermittent preventive treatment for malaria during ANC, and receiving a neonatal tetanus vaccination during ANC; *E*_*ijk*_ is whether the woman completed primary education or not; *W*_*jk*_ is whether or not house *j* is rich or poor; and *S*_*k*_ is the survey dataset.

### Statistical methods

We modeled neonatal mortality as a function of receiving deworming medicine during ANC after adjusting for the following *a priori* determined covariates: the mother’s age (categorized as < 18, 18-35, and >35), the mother’s parity and birth order (categorized as firstborn, 2^nd^ or 3^rd^ born with < 24 months preceding birth interval, 2^nd^ or 3^rd^ born with ≥ 24 months preceding birth interval, 4^th^ or later born with < 24 months preceding birth interval, and 4^th^ or later born with ≥ 24 months preceding birth interval), the presence of a skilled birth attendant during childbirth (doctor, nurse, or midwife), the location of child birth (at a health center or not), the household wealth quintile, the mother’s education (categorized as no education, some primary, or completed primary or higher), the location of the house (urban or rural), the household’s sanitation access (categorized as any or none), and the survey dataset. We utilized a Poisson model with the number of days alive (up to 28) included as the exposure and the matched group included as a random intercept. The analysis of neonatal mortality can be described using the following equation:

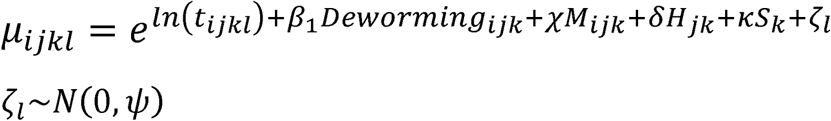

where *µ*_*ijkl*_ is the rate of death for child *i* in house *j* in survey *k* in matched group *l, t*_*ijkl*_ is the number of days the child was alive or at risk of death, *Deworming*_*ijk*_ is whether or not the child’s mother received deworming during ANC, *M*_*ijk*_ is a vector of a child’s mother’s characteristics, *H*_*jk*_ is a vector a child’s household’s characteristics,*S*_*k*_ is a vector of survey characteristics and *ζ*_*l*_ is a random intercept for matched group *l* that is assumed to be normally distributed with a mean of zero.

We modeled low birth weight as a function of receiving deworming medicine during ANC after adjusting for the same *a priori* determined covariates as previously described. We utilized a logit model with the matched group included as a random intercept. The analysis of low birth weight can be described using the following equation:

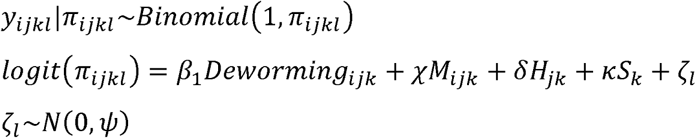

where *π*_*ijkl*_ is a dichotomous outcome for child *i* in household *j* in survey *k* in matched group *l, Deworming*_*ijk*_ is whether the child’s mother received deworming medicine during ANC, *M*_*ijk*_ is a vector of a child’s mother’s characteristics, *H*_*jk*_ is a vector of a child’s household’s characteristics, is a vector of survey characteristics and *ζ*_*l*_ is a random intercept for matched group *l* that is assumed to be normally distributed with a mean of zero. All analyses were conducted in Stata version 15.1.

We also tested whether the association between deworming medicine and the outcomes of neonatal mortality and low birthweight was moderated by the prevalence of any STH in the country. Based upon estimates provided by Pullan et al. [17], we categorized countries as being low (< 20%) or high (> 20%) STH prevalence and then tested for an interaction between low/high STH prevalence and receiving deworming during ANC using a likelihood ratio test of the log likelihoods.

## Findings

### Participants

As of October 2019, a total of 290 survey datasets listed at www.dhsprogram.com contained information on either neonatal mortality, low birthweight, or deworming during ANC. Deworming during ANC was available for 95 of these datasets. One survey dataset (Rwanda 2007-08) did not have measures of low birth weight. This left 95 datasets available to create a retrospective cohort of 825,492 single live births to assess the impact of deworming during ANC on neonatal mortality and 94 datasets available to create a retrospective cohort of 807,957 single live births to assess the impact of deworming during ANC on low birthweight. Following exact matching, 95 datasets and 797,772 women were available for the outcome of neonatal mortality and 94 datasets and 772,155 women were available for the outcome of low birthweight. Figure 1 shows the geographic distribution of countries included in the analyses.

### Descriptive data

Among matched births, 25% of mothers reported receiving deworming medicine during ANC. Two percent of births resulted in a neonatal death (n = 15,784). Only 61% (n = 487,763) of mothers reported a measured birthweight. Among births with a measured birthweight, 12.5% (n = 61,177) were < 2500 grams. Mean birthweight was 3,072g (standard deviation = 691g). Among 770,300 births with a perceived birth size, 93,802 mothers (12%) reported the baby to be “smaller than average” and 39,601 mothers reported the baby to be “very small” (5%). Table 1 provides dataset-level information on deworming during ANC and birth outcomes.

**Table 1:** Number of women receiving deworming medicine during routine ANC and birth outcomes.

| Survey dataset | Coverage of deworming during ANC | Neonatal deaths without deworming / N without deworming | Neonatal deaths with deworming / N with deworming | LBW without deworming / N without deworming | LBW with deworming / N with deworming |
| --- | --- | --- | --- | --- | --- |
| **Afghanistan 2015** | 3% | 396 / 17,773 | 20 / 616 | 3,800 / 17,355 | 136 / 605 |
| **Albania 2008-09** | 3% | 6 / 933 | 0 / 31 | 30 / 930 | 0 / 31 |
| **Albania 2017-18** | 2% | 2 / 1952 | 0 / 45 | 79 / 1,950 | 2 / 45 |
| **Angola 2015-16** | 46% | 113 / 4,607 | 69 / 3,939 | 514 / 4,262 | 365 / 3,855 |
| **Armenia 2010** | 1% | 5 / 1,136 | 0 / 7 | 70 / 1,134 | 1 / 7 |
| **Azerbaijan 2006** | 3% | 31 / 1,430 | 1 / 51 | 123 / 1,201 | 8 / 47 |
| **Benin 2011-12** | 76% | 44 / 2,066 | 97 / 6,397 | 282 / 1,829 | 766 / 6,205 |
| **Benin 2017-18** | 66% | 66 / 2,841 | 122 / 5,578 | 379 / 2,801 | 616 / 5,529 |
| **Burkina Faso 2010** | 26% | 138 / 7,471 | 32 / 2,562 | 1,025 / 7,456 | 299 / 2,559 |
| **Burundi 2010-11** | 31% | 85 / 3,339 | 29 / 1,1519 | 445 / 3,281 | 155 / 1,503 |
| **Burundi 2016-17** | 66% | 31 / 2,888 | 93 / 5,708 | 361 / 2,879 | 582 / 5,697 |
| **Cambodia 2005** | 12% | 125 / 4,979 | 9 / 700 | 740 / 4,940 | 84 / 692 |
| **Cambodia 2010** | 50% | 74 / 3,105 | 45 / 3,106 | 344 / 2,969 | 226 / 3,051 |
| **Cambodia 2014** | 74% | 24 / 1,442 | 47 / 4,212 | 157 / 1,428 | 330 / 4,200 |
| **Cameroon 2011** | 43% | 86 / 4,090 | 58 / 3,069 | 579 / 4,059 | 299 / 3,053 |
| **Chad 2014-15** | 23% | 185 / 8,076 | 44 / 2,382 | 2,362 / 8,011 | 508 / 2,376 |
| **Comoros 2012** | 60% | 19 / 722 | 18 / 1,077 | 178 / 685 | 209 / 1,048 |
| **Congo 2011-12** | 81% | 22 / 1,167 | 71 / 4,888 | 145 / 1,128 | 469 / 4,858 |
| **Cote d'Ivoire 2011** | 38% | 96 / 3,110 | 43 / 1,877 | 447 / 3,024 | 271 / 1,851 |
| **DRC 2013-14** | 52% | 121 / 5,190 | 107 / 5,724 | 451 / 5,075 | 470 / 5,667 |
| **Dominican Republic 2013** | 13% | 41 / 2,478 | 4 / 354 | 345 / 2,472 | 46 / 354 |
| **Egypt 2014** | 4% | 85 / 9,981 | 3 / 397 | 1,413 / 9,968 | 51 / 393 |
| **Eswatini 2006-07** | 12% | 37 / 1,583 | 5 / 212 | 121 / 1,557 | 15 / 207 |
| **Ethiopia 2011** | 6% | 176 / 7,099 | 15 / 422 | 2,283 / 7,081 | 120 / 422 |
| **Ethiopia 2016** | 6% | 149 / 6,438 | 10 / 415 | 1,668 / 6,366 | 95 / 411 |
| **Gabon 2012** | 64% | 27 / 1,416 | 44 / 2,495 | 218 / 1,302 | 323 / 2,434 |
| **Gambia 2013** | 43% | 38 / 2,947 | 27 / 2,215 | 541 / 2,939 | 300 / 2,211 |
| **Ghana 2008** | 40% | 28 / 1,139 | 14 / 746 | 136 / 1,130 | 90 / 743 |
| **Ghana 2014** | 44% | 41 / 2,280 | 31 / 1,800 | 1,203 / 8,431 | 89 / 623 |
| **Guatemala 2014-15** | 7% | 110 / 8,438 | 8 / 623 | 1,203 / 8,431 | 89 / 623 |
| **Guinea 2012** | 30% | 88 / 3,368 | 23 / 1,473 | 387 / 3,361 | 131 / 1,473 |
| **Guyana 2009** | 20% | 17 / 1,135 | 4 / 289 | 175 / 1,127 | 41 / 285 |
| **Haiti 2005-06** | 7% | 74 / 3,566 | 9 / 286 | 1,083 / 3,566 | 78 / 286 |
| **Haiti 2012** | 15% | 104 / 4,294 | 16 / 785 | 1,331 / 4,288 | 199 / 785 |
| **Haiti 2016-17** | 10% | 100 / 4,102 | 7 / 477 | 991 / 4,102 | 99 / 477 |
| **Honduras 2005-06** | 8% | 78 / 6,632 | 11 / 547 | 1,101 / 6,625 | 108 / 544 |
| **Honduras 2011-12** | 7% | 92 / 7,687 | 12 / 612 | 1,069 / 7,681 | 97 / 612 |
| **India 2005-06** | 4% | 782 / 34,248 | 26 / 1,433 | 7,304 / 33,761 | 284 / 1,422 |
| **India 2015-16** | 15% | 3,504 / 159,219 | 468 / 29,185 | 25,971 / 155,748 | 4,785 / 28,940 |
| **Kenya 2008** | 20% | 57 / 2,770 | 13 / 672 | 348 / 2,747 | 54 / 668 |
| **Kenya 2014** | 31% | 66 / 4,744 | 41 / 2,179 | 549 / 4,679 | 202 / 2,172 |
| **Liberia 2007** | 30% | 62 / 2,600 | 20 / 1,108 | 539 / 2,592 | 180 / 1,106 |
| **Liberia 2013** | 57% | 55 / 2,230 | 61 / 2,978 | 459 / 2,228 | 543 / 2,971 |
| **Madagascar 2008-09** | 40% | 93 / 4,984 | 47 / 3,373 | 916 / 4,911 | 531 / 3,320 |
| **Malawi 2010** | 28% | 209 / 9,518 | 78 / 3,759 | 1,176 / 9,375 | 469 / 3,706 |
| **Malawi 2015-16** | 52% | 123 / 6,289 | 115 / 6,816 | 780 / 6,245 | 841 / 6,781 |
| **Maldives 2009** | 18% | 17 / 2,482 | 0 / 552 | 261 / 2,481 | 61 / 552 |
| **Mali 2012-13** | 29% | 120 / 4,461 | 42 / 1,847 | 732 / 4,225 | 225 / 1,818 |
| **Mali 2018** | 49% | 83 / 3,108 | 60 / 2,941 | 524 / 2,787 | 571 / 2,875 |
| **Mozambique 2011** | 33% | 137 / 4,804 | 66 / 2,390 | 593 / 4,594 | 286 / 2,352 |
| **Myanmar 2015-16** | 57% | 33 / 1,614 | 32 / 2,127 | 214 / 1,531 | 223 / 2,079 |
| **Namibia 2006-07** | 8% | 58 / 3,293 | 6 / 289 | 460 / 3,261 | 48 / 281 |
| **Namibia 2013** | 7% | 52 / 3,337 | 4 / 265 | 439 / 3,310 | 34 / 262 |
| **Nepal 2006** | 19% | 80 / 3,276 | 15 / 786 | 639 / 3,274 | 139 / 786 |
| **Nepal 2011** | 59% | 33 / 1,629 | 42 / 2,353 | 302 / 1,627 | 379 / 2,352 |
| **Nepal 2016** | 74% | 21 / 1,021 | 28 / 2,910 | 163 / 1,020 | 402 / 2,905 |
| **Niger 2012** | 51% | 64 / 3,705 | 65 / 3,793 | 854 / 3,490 | 810 / 3,748 |
| **Nigeria 2008** | 10% | 440 / 15,188 | 36 / 1,656 | 2,379 / 15,062 | 172 / 1,650 |
| **Nigeria 2013** | 16% | 465 / 16,073 | 91 / 3,001 | 2,445 / 15,971 | 315 / 2,991 |
| **Pakistan 2012-13** | 2% | 235 / 6,136 | 5 / 156 | 1,340 / 6,123 | 38 / 156 |
| **Pakistan 2017-18** | 2% | 194 / 6,823 | 2 / 163 | 1,344 / 6,804 | 32 / 159 |
| **Peru 2004-08** | 4% | 79 / 8,864 | 1 / 348 | 898 / 8,858 | 38 / 348 |
| **Peru 2009** | 3% | 72 / 7,871 | 1 / 280 | 750 / 7,866 | 30 / 280 |
| **Peru 2010** | 3% | 59 / 7,056 | 1 / 253 | 687 / 7,055 | 32 / 253 |
| **Peru 2011** | 3% | 60 / 6,967 | 2 / 251 | 646 / 6,966 | 28 / 251 |
| **Peru 2012** | 3% | 60 / 6,887 | 1 / 243 | 576 / 6,84 | 28 / 243 |
| **Philippines 2008** | 5% | 45 / 4,011 | 3 / 196 | 823 / 3,999 | 54 / 196 |
| **Philippines 2013** | 5% | 44 / 4,571 | 5 / 263 | 782 / 3,719 | 48 / 214 |
| **Philippines 2017** | 7% | 85 / 7,016 | 5 / 498 | 837 / 5,939 | 68 / 423 |
| **Rwanda 2007-08** | 19% | 43 / 2,567 | 9 / 617 | . | . |
| **Rwanda 2010** | 40% | 62 / 3,754 | 34 / 2,453 | 348 / 3,742 | 178 / 2,443 |
| **Rwanda 2014-15** | 50% | 44 / 2,934 | 37 / 2,945 | 217 / 2,923 | 179 / 2,934 |
| **STP 2008** | 59% | 6 / 551 | 7 / 794 | 45 / 537 | 52 / 770 |
| **Senegal 2010-11** | 25% | 141 / 5,656 | 51 / 1,927 | 1,143 / 5,641 | 388 / 1,922 |
| **Senegal 2012-13** | 28% | 59 / 2,823 | 17 / 1,107 | 697 / 2,821 | 236 / 1,105 |
| **Senegal 2014** | 30% | 110 / 5,664 | 32 / 2,411 | 1,406 / 5,662 | 512 / 2,409 |
| **Senegal 2015** | 32% | 54 / 2,903 | 31 / 1,369 | 672 / 2,902 | 234 / 1,368 |
| **Senegal 2016** | 35% | 66 / 2,698 | 19 / 1,466 | 642 / 2,697 | 241 / 1,462 |
| **Senegal 2017** | 44% | 107 / 4,290 | 72 / 3,380 | 891 / 4,286 | 670 / 3,374 |
| **Sierra Leone 2008** | 47% | 61 / 1,945 | 57 / 1,720 | 325 / 1,913 | 283 / 1,687 |
| **Sierra Leone 2013** | 74% | 79 / 2,110 | 198 / 6,140 | 326 / 2,050 | 675 / 6,038 |
| **Tajikistan 2017** | 2% | 45 / 4,000 | 0 / 83 | 296 / 3,867 | 10 / 79 |
| **Tanzania 2015-16** | 63% | 47 / 2,563 | 74 / 4,306 | 273 / 2,542 | 362 / 4,284 |
| **Timor-Leste 2009-10** | 14% | 97 / 4,918 | 10 / 832 | 727 / 4,813 | 110 / 831 |
| **Timor-Leste 2016** | 18% | 61 / 3,931 | 19 / 840 | 358 / 3,252 | 60 / 763 |
| **Togo 2013-14** | 62% | 38 / 1,807 | 57 / 2,920 | 287 / 1,789 | 292 / 2,904 |
| **Uganda 2006** | 27% | 63 / 3,464 | 23 / 1,293 | 648 / 3,435 | 203 / 1,289 |
| **Uganda 2011** | 51% | 69 / 2,326 | 39 / 2,375 | 414 / 2,281 | 324 / 2,321 |
| **Uganda 2016** | 61% | 112 / 3,888 | 110 / 6,129 | 574 / 3,800 | 692 / 6,050 |
| **Ukraine 2007** | 7% | 4 / 960 | 1 / 75 | 33 / 960 | 7 / 75 |
| **Yemen 2013** | 3% | 174 / 9,776 | 8 / 352 | 3,109 / 9,756 | 137 / 351 |
| **Zambia 2007** | 39% | 69 / 2,293 | 31 / 1,450 | 237 / 2,273 | 109 / 1,444 |
| **Zambia 2013** | 66% | 56 / 3,038 | 84 / 6,004 | 335 / 3,008 | 527 / 5,941 |
| **Zimbabwe 2010-11** | 3% | 59 / 3,211 | 1 / 103 | 279 / 3,164 | 10 / 103 |
| **Zimbabwe 2015** | 3% | 63 / 4,152 | 3 / 145 | 391 / 4,145 | 9 / 145 |

### Main results

Before adjusting for any other covariates, 2.1% of births where mothers did not receive deworming during ANC (12,873 / 610,118) and 1.8% of births where mothers did receive deworming (3,622 / 202,501) during ANC died within the neonatal period (relative risk of cumulative incidence = 0.85). Before adjusting for any other covariates, 16.9% of babies where mothers did not receive deworming during ANC (102,802 / 505,383) and 13.3% babies where mothers did receive deworming during ANC (26,590 / 199,772) were considered low birth weight (relative risk of cumulative incidence = 0.79).

After adjusting for selection bias via exact matching and including other factors hypothesized to be associated with neonatal mortality, receiving deworming during routine ANC was associated with a 14% reduction in the risk of neonatal mortality (IRR = 0.86, 95% CI = 0.82 – 0.90) (Table 2). This relationship was not moderated by STH prevalence: the likelihood ratio test for interaction was not statistically significant (likelihood ratio [LR] = 2.19, p = 0.139).

**Table 2:** Results from Poisson regression analysis of deworming during ANC and neonatal mortality.

| Measure | Categorization | Unadjusted IRR<br>(95% CI) | P-value | Adjusted IRR<br>(95% CI) | P-value |
| --- | --- | --- | --- | --- | --- |
| **Deworming** | No deworming during ANC | Ref. | Ref. | Ref. | Ref. |
|  | Deworming during ANC | 0.81 (0.78 – 0.85) | < 0.001 | 0.86 (0.82 – 0.89) | < 0.001 |
| **Skilled birth attendant** | No doctor, nurse, or midwife during delivery | Ref. | Ref. | Ref. | Ref. |
|  | Doctor, nurse, or midwife during delivery | 0.91 (0.88 – 0.94) | < 0.001 | 1.01 (0.94 – 1.08) | 0.767 |
| **Facility delivery** | Child was delivered somewhere other than a health center | Ref. | Ref. | Ref. | Ref. |
|  | Child was delivered at a health center | 0.92 (0.89 – 0.95) | < 0.001 | 1.11 (1.04 – 1.19) | 0.002 |
| **Mother's age at delivery** | 18-35 years old | Ref. | Ref. | Ref. | Ref. |
|  | < 20 years old | 1.31 (1.25 – 1.38) | < 0.001 | 1.18 (1.11 – 1.24) | < 0.001 |
|  | > 35 years old | 1.59 (1.52 – 1.66) | < 0.001 | 1.59 (1.52 – 1.66) | < 0.001 |
| **Mother's parity** | Child is firstborn | Ref. | Ref. | Ref. | Ref. |
|  | 2<sup>nd</sup> born ≥ 24 month preceding birth interval | 0.67 (0.64 – 0.71) | < 0.001 | 0.69 (0.65 – 0.73) | < 0.001 |
|  | 2<sup>nd</sup> born < 24 month preceding birth interval | 0.91 (0.85 – 0.98) | 0.013 | 0.91 (0.85 – 0.98) | 0.002 |
|  | 3<sup>rd</sup> born or later ≥ 24 month preceding birth interval | 0.90 (0.86 – 0.93) | < 0.001 | 0.78 (0.74 – 0.82) | < 0.001 |
|  | 3<sup>rd</sup> born or later < 24 month preceding birth interval | 1.45 (1.38 – 1.52) | < 0.001 | 1.30 (1.23 – 1.38) | < 0.001 |
| **Mother's education** | No education | Ref. | Ref. | Ref. | Ref. |
|  | Some primary school | 0.95 (0.90 – 0.99) | 0.022 | 1.02 (0.97 – 1.07) | 0.488 |
|  | Completed primary school | 0.93 (0.88 – 0.99) | 0.020 | 1.02 (0.95 – 1.10) | 0.529 |
|  | Some secondary school or higher | 0.71 (0.68 – 0.74) | < 0.001 | 0.87 (0.82 – 0.92) | < 0.001 |
| **Household location** | Rural | Ref. | Ref. | Ref. | Ref. |
|  | Urban | 1.14 (1.10 – 1.18) | < 0.001 | 0.97 (0.93 – 1.01) | 0.139 |
| **Sanitation access** | Any type of sanitation access | Ref. | Ref. | Ref. | Ref. |
|  | No sanitation access | 1.29 (1.24 – 1.33) | < 0.001 | 1.13 (1.09 – 1.18) | < 0.001 |
| **Household wealth** | Poorest wealth quintile | Ref. | Ref. | Ref. | Ref. |
|  | Poorer wealth quintile | 0.98 (0.94 – 1.03) | 0.403 | 1.04 (0.99 – 1.09) | 0.120 |
|  | Middle wealth quintile | 0.90 (0.86 – 0.94) | < 0.001 | 1.05 (0.99 – 1.10) | 0.105 |
|  | Richer wealth quintile | 0.82 (0.78 – 0.86) | < 0.001 | 1.01 (0.95 – 1.08) | 0.743 |
|  | Richest wealth quintile | 0.69 (0.66 – 0.73) | < 0.001 | 0.92 (0.85 – 0.99) | 0.024 |

N = 797,771 children, 7,255 matched groups (adjusted model only), 95 survey datasets ranging from year 2005-2018. Unadjusted model standard errors were adjusted for survey cluster and included survey
dataset as a covariate. Adjusted model included a random intercept of matched group and survey dataset as a covariate.

After adjusting for selection bias via exact matching and including other factors hypothesized to be associated with low birthweight, receiving deworming during routine ANC was associated with a 6% reduction in the odds of low birthweight (Odds ratio [OR] = 0.94, 95% CI = 0.92 – 0.95). This relationship was moderated by STH prevalence (LR = 23.87, p < 0.001). In low transmission countries (< 20% national STH prevalence according to Pullan et al. [17]), deworming during routine ANC was associated with an 11% reduction in the odds of low birthweight (OR = 0.89, 95% CI = 0.87 – 0.92). In high transmission countries (> 20% national STH prevalence), deworming during routine ANC was associated with a 3% reduction in the odds of low birthweight (OR = 0.97, 95% CI = 0.95 – 0.99). Table 3 shows the results from the unadjusted and adjusted analyses of low birthweight.

**Table 3:**
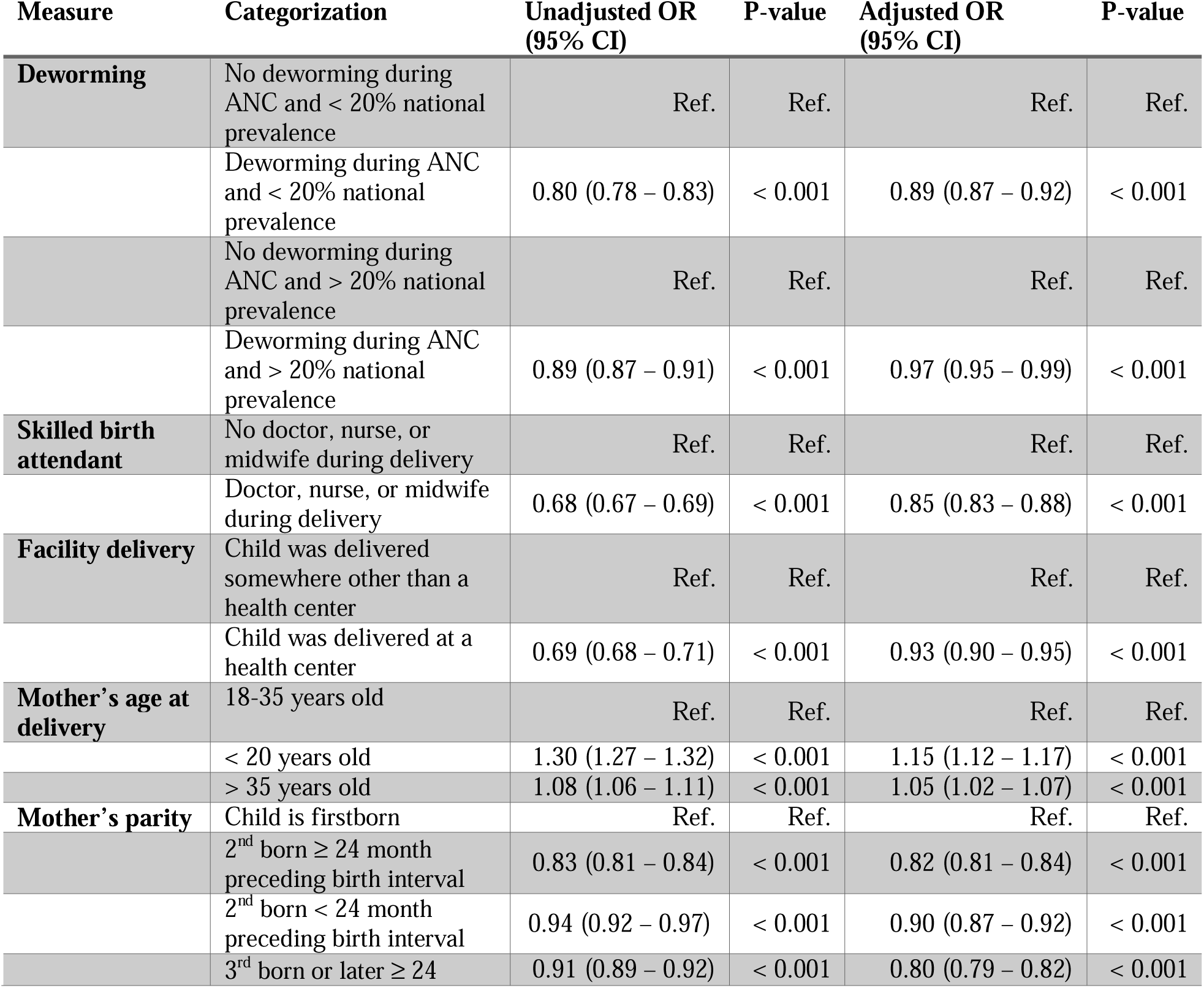
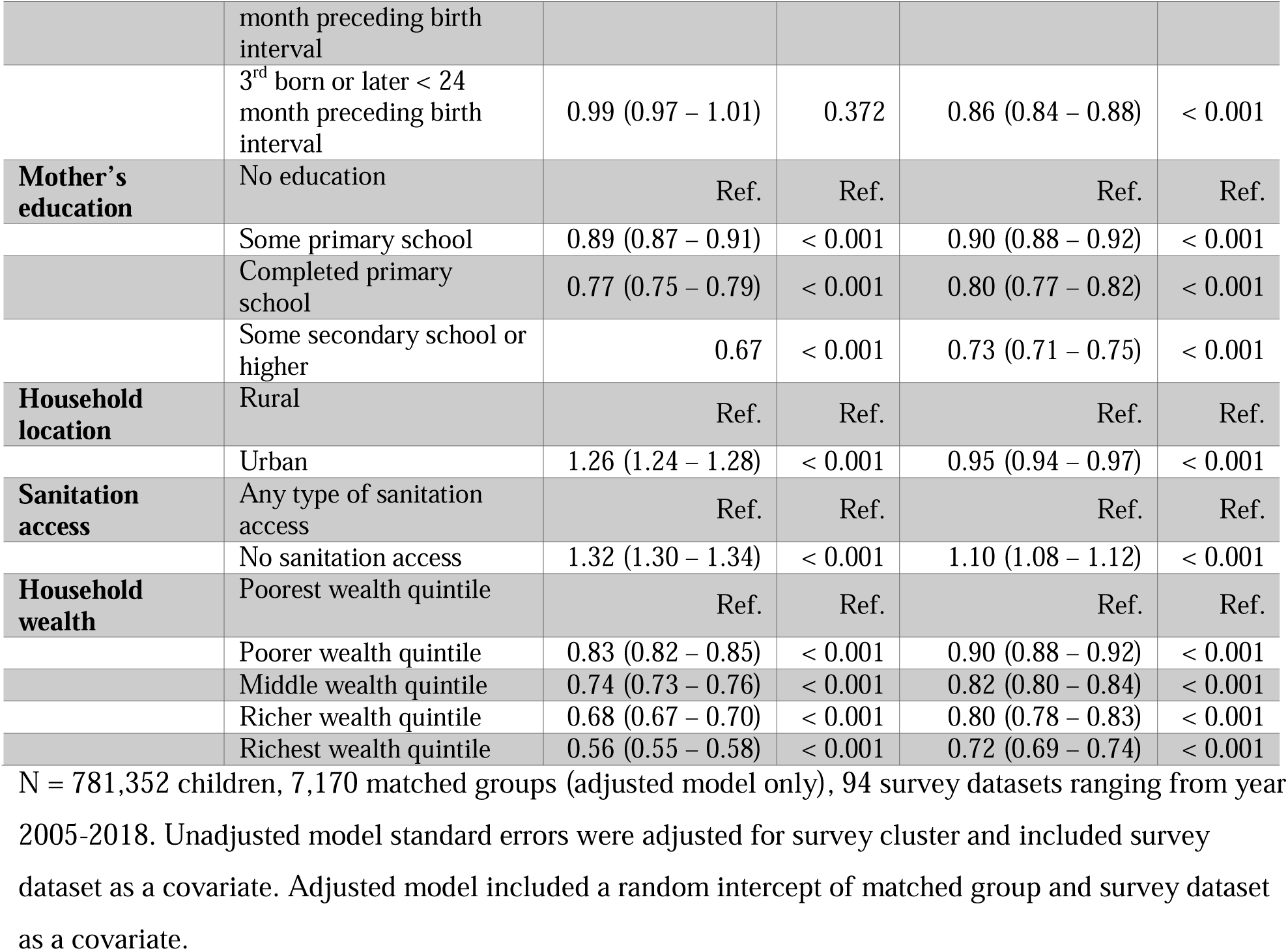
Results from logistic regression analysis of deworming during ANC and low birthweight.

Among the children who were weighed at birth, the effect of deworming during ANC on low birthweight was somewhat smaller (OR = 0.97, 95% CI = 0.95 – 1.00, p = 0.022). There was also a significant interaction between STH prevalence and receipt of deworming medicine during ANC (LR 10.28, p = 0.0013). Deworming during ANC effectively reduced the odds of low birth weight in low transmission countries (OR = 0.93, 95% CI = 0.89 – 0.96), but not in high transmission countries (OR = 1.00, 95% CI = 0.97 – 1.03).

## Interpretation

### Key results

We provide empirical evidence as to the impact of deworming in pregnant women on the neonatal health of their infants (low birthweight and neonatal mortality) across a large range of STH endemic countries. Using a retrospective birth cohort of more than 770,000 births, we find that children born to mothers who received deworming during antenatal care have a 14% lower risk of neonatal mortality (95% CI = 11 – 16%). In countries with < 20% estimated national prevalence of any STH, deworming during antenatal care was associated with an 11% reduction in the odds of low birthweight (95% CI = 8 – 13%). In countries with > 20% estimated national prevalence of any STH, deworming during ANC was associated with a 3% reduction in the odds of low birthweight (95% CI = 1-5%).

### Limitations

A number of limitations are present in this analysis. First, there is selection bias in women receiving deworming during ANC. Due to the inequity observed in children receiving deworming medicines [18], we suspect that mothers receiving deworming during ANC are predisposed to have better birth outcomes than mothers not receiving deworming during ANC. We have attempted to mitigate the selection bias by matching mothers on the probability that they receive deworming during ANC before analysis. There may still be some residual confounding in this analysis. Second, this study relies on mothers’ recall of medicines received during their most recent pregnancy. There may be some recall error associated with mothers’ abilities to remember all the medicines and supplements they received during pregnancy, and recall bias may be present if mothers of babies suffering neonatal deaths are more likely remember receiving deworming medicine or not better than mothers of surviving babies. We have attempted to mitigate this limitation by only including the most recent pregnancy and only including pregnancies within 5 years of the study. Third, because this study creates a retrospective cohort from historical birth outcomes we are unable to account for the mother’s or child’s health status at the time of pregnancy and birth. Fourth, this study relies on mothers’ recall of the age of the child at death, which may lead to misclassification error in our outcome of neonatal mortality. As such, we observed heaping of death at one month of age. We have chosen to include children dying at one month of age as neonatal deaths. We do not expect the misclassification error to be related to receiving deworming during ANC and suspect no bias in this regard. And fifth, our measure of low birthweight is a composite indicator of a mother’s perception of birth size or a mother’s recall of the child’s birth weight if the child was weighed at birth. This approach may lead to misclassification error in our outcome of low birth weight, but we do not expect it to bias the results as it is not likely to be associated with receiving deworming medicine during ANC. There were also no differences in the results when we limited the analysis to those babies who were weighed at birth.

### Interpretation

These results suggest a strong benefit of deworming during ANC that is perhaps even more pronounced in low transmission areas. The half-life of deworming medicines is quite short (< 24 hours), and very little chemoprophylaxis is provided for women treated. Reinfection occurs rapidly in areas of high transmission, particularly among those who were previously infected [19,20]. Deworming should only be considered a stopgap measure, useful to reduce harm from STH infections as the world pursues universal access to improved sanitation. However, deworming should be available to all pregnant women in STH-endemic countries, given the increased risk of low birthweight and neonatal mortality when living in communities where access to sanitation is limited [21].

Periodic deworming is a very low-cost intervention. Where infrastructure for distribution is in place, such as through school programs, vaccination campaigns or routine ANC, the intervention cost is a few cents for each individual treated [22,23]. Considered safe during pregnancy, the risk of side effects from the drug administration is also minimal because benzimidazoles are poorly absorbed and are normally expelled after killing the worms present in the intestine [24]. These results provide impetus to improve access to deworming medicines during routine ANC.

While some have called for randomized trials of deworming for pregnant women to establish efficacy, these results suggest that deworming reduces the risk of neonatal mortality and low birthweight. To our knowledge this is the largest analysis conducted on deworming and birth outcomes (more than 770,000 births – previous Cocherane review has < 3,400), and in our view is the more appropriate approach to provide evidence on a relatively rare event like neonatal mortality. From an empirical perspective, a sample size of 3,400 does not allow for a great deal of variation in the outcome variable. That is, for low probability (*p*) events, *np* is typically low *n* unless is sufficintly large to compensate. The approach we used herein also avoids the ethical dilemma of withholding an intervention from the control group, the major limitation to conducting randomized control trials of clearing parasites with deworming medicine.

The WHO recommends periodic deworming of children and women of reproductive age (including pregnant women after the first trimester) [24]. While in the last 10 years the coverage of the intervention scaled up significantly for preschool and school age children, reaching over 65% in 2017 [25], the scale up of coverage for women of reproductive age has been much slower, with an average of 23% of pregnant women in STH endemic countries receiving deworming during ANC [26]. A recent meeting of the WHO Advisory Group on deworming in girls and women of reproductive age [27] urged all stakeholders in women’s health to take immediate action in their respective domains to ensure that women of reproductive age are now included in their STH policies and programs, and to invite WHO to develop support material to facilitate implementation of deworming programs [28].

The traditional understanding of host-parasite interactions during pregnancy is that the parasite takes nutrients from the pregnant host, that either causes anemia and/or reduces the nutrients passing to the fetus leading to intrauterine growth restriction [29,30], increased risk of low birthweight, and then subsequent neonatal mortality [31]. In this analysis, the impact of deworming on low birthweight was quite modest in high transmission areas (3% reduction) compared to the impact of deworming on neonatal mortality (14% reduction). These results provide some evidence that helminth infections may indeed decrease nutrient flow to the fetus, but perhaps raise questions about other ways in which helminth infections affect pregnancy.

We suspect the primary mechanism of protection for deworming medicines to be through improving the health of the mother and subsequently the health of the fetus. In order to survive within their hosts, STH modify their hosts’ immune responses, downregulating T-cell activity and other immune responses [32]. This immunosuppression affects not only the hosts’ immune response to the STH, but also the immune response to other pathogens. For example, chronic STH infection is associated with suboptimal immunity following the receipt of various vaccines [33]. And, in areas of high prevalence of STH, routine deworming led to improved immune responses to non-STH pathogens [34]. This effect of compromising maternal immunity may be particularly serious given that a pregnant woman’s immune response is downregulated by the 18^th^ week of fetal gestation to ensure the mother’s body does not reject the fetus as a foreign body [35,36]. Furthermore, immune system downregulation passes through to the fetus. Helminth infections prime the immune system of the fetus, making children born to mothers with helminth infections more vulnerable to not only helminth infections after they are born, but other pathogens as well [37,38]. This mechanism could lead to increased risk of neonatal mortality for children of women who do not receive deworming medicines during antenatal care.

### Conclusion

This large retrospective cohort of survey data suggests that deworming during antenatal care provides a protective benefit against neonatal mortality and low birthweight. This protection is even greater in countries with lower STH transmission.

## Data Availability

All data utilized in this manuscript are publicly available from the Demographic and Health Surveys program

https://dhsprogram.com

## Other information

### Author contributions

Conceptualization, DAL; Methodology, BW, BK, DAL; Formal analysis, DAL; Data curation, BK, DAL; Writing – original draft, BW, DAL; Writing – review and editing, BW, BK, SL, TE, AM DAL; Visualization, DAL; Supervision, BK, DAL; Project administration, BK, DAL.

## Acronyms

ANC: antenatal care
CI: confidence interval
DHS: demographic health survey
IRR: incident rate ratio
LR: likelihood ratio
OR: odds ratio

